# Recessive loss-of-function and dominant gain-of-function variants in *FBXO31* are associated with distinct neurodevelopmental disorders

**DOI:** 10.64898/2026.09.24.26363382

**Authors:** Jonas Böhnlein, Birgit Weiss, Steffen Hirsch, Johann G. Maass, Katharina Obeid, Katrin Hinderhofer, Sebastian Schröder, Björn Sämann, Maja Hempel, Sebastian Sailer

## Abstract

*FBXO31* encodes the substrate receptor of an SCF (SKP1-CUL1-F-box) E3 ubiquitin ligase that recognizes C-terminal amide-bearing proteins (CTAPs) and targets them for proteasomal degradation, a function of particular importance in neurons. A recurrent *de novo* missense variant, c.1000G>A p.(Asp334Asn), reprograms *FBXO31* substrate specificity and causes an autosomal-dominant neurodevelopmental disorder with spastic cerebral palsy through a neomorphic gain-of-function mechanism. By contrast, biallelic loss of function has tentatively been linked to a recessive disorder. Here we report a female patient, born to consanguineous parents, with moderate intellectual disability, developmental delay and dysmorphic features, who carries a homozygous ~54 kb deletion removing exons 2-8 and the coding part of exon 9 of *FBXO31*. The deletion retains only the exon 1-encoded F-box domain, while removing the coding sequence for the entire C-terminal substrate-recognition region. It is predicted to result in loss of function. Segregation is consistent with autosomal-recessive inheritance, with both parents and three of five unaffected siblings being heterozygous carriers. Together with two previously reported recessive families and the experimentally established neomorphic action of p.(Asp334Asn), the homozygous deletion supports an *FBXO31* allelic series comprising dominant gain of function and recessive biallelic loss of function.

## Introduction

Intellectual disability (ID) is a neurodevelopmental disorder marked by impaired intellectual and adaptive functioning (1). Genetically, ID is highly heterogeneous, with more than 1,500 genes implicated across X-linked, autosomal-dominant and autosomal-recessive forms (2, 3).

*FBXO31* encodes an F-box protein that functions as the substrate-recognition component of the SCF-FBXO31 E3 ubiquitin ligase and was previously linked to cell-cycle regulation after DNA damage (4, 5). More recently, *FBXO31* was shown to recognize C-terminal amide-bearing proteins (CTAPs) and target them for proteasomal degradation, a proteostasis pathway of particular relevance in mature neurons (6). The recurrent *de novo* missense variant NM_024735.5:c.1000G>A p.(Asp334Asn) disrupts CTAP recognition and reprograms substrate specificity via a neomorphic gain-of-function mechanism (6). Clinically, this variant causes an autosomal-dominant neurodevelopmental disorder with spastic cerebral palsy, speech impairment and intellectual disability, recently delineated as Kruer syndrome (7-9).

In contrast, the consequences of *FBXO31* loss of function remain incompletely defined. Previously reported heterozygous deletions encompassing *FBXO31* include multiple other genes, preventing confident attribution of their associated neurodevelopmental and congenital features to monoallelic loss of *FBXO31* alone (10-14).

Evidence linking biallelic *FBXO31* loss of function to human disease is limited. Two studies described homozygous *FBXO31* variants associated with autosomal-recessive intellectual developmental disorder 45 (MRT45; OMIM #615979): a deletion-insertion frameshift, p.(Cys283Asnfs*81), predicted to truncate FBXO31 and associated with reduced transcript and protein abundance, and a missense variant, p.(Arg511Gln), supported by segregation and *in silico* predictions but lacking functional validation. Affected individuals had intellectual disability and variable facial dysmorphism without spasticity or other motor involvement (15, 16). Here we report a homozygous, intragenic *FBXO31* deletion that removes the coding sequence for the C-terminal substrate-recognition region and is predicted to result in loss of function. We contextualize this finding using population and clinical variation data and recent mechanistic insights into *FBXO31* biology. Together with previously reported biallelic *FBXO31* variants, our findings strengthen the evidence for a recessive loss-of-function mechanism distinct from the dominant gain-of-function mechanism underlying Kruer syndrome.

## Materials and Methods

### Patient recruitment and ethics

Our patient was enrolled at the University Hospital Heidelberg, Germany, and underwent genetic diagnostics to identify the molecular cause of her neurodevelopmental disorder. Patient and family enrollment was approved by the local ethics committee (Medical Faculty of Heidelberg University, S-632/2023). Written informed consent for genetic testing and for publication was obtained from all participating family members or, for minors, from their legal guardians.

### DNA microarray

Genomic DNA from peripheral blood leukocytes was hybridized onto the Affymetrix CytoScan HD Oligo/SNP Array (Affymetrix, Santa Clara, CA, USA; 2.67 million markers; average marker spacing 1,148 bp, genome-wide). Arrays were analyzed with Chromosome Analysis Suite v4.2.1 and Annotation NetAffx Build 20210602 (June 2021). Annotations refer to the human reference sequence GRCh37. Thresholds for detection were 25 markers and a minimum size of 50 kb for deletions and gains; long contiguous stretches of homozygosity were reported from 10 Mb. Detected imbalances were evaluated against control datasets embedded in the software and resources including DGV, ClinGen, ClinVar, DECIPHER, OMIM and the UCSC Genome Browser. Variants were classified following ACMG/ClinGen standards (17).

### Exome sequencing

Coding regions and adjacent intronic sequences were enriched using the SureSelect Human All Exon V7 kit (Agilent) and sequenced as 100 bp paired-end reads on a NovaSeq 6000 platform (Illumina, San Diego, CA, USA). Bioinformatic analysis was performed using the varfeed pipeline with annotation on GRCh38/hg38. Coverage was evaluated per exon, and regions below 10× average depth were excluded from sequence-variant interpretation. Rare coding and splice-region SNVs/indels were prioritized according to allele frequency, predicted functional impact, phenotype compatibility and inheritance pattern. Population frequencies were derived from gnomAD and internal databases, and variants were classified according to ACMG/AMP guidelines.

Because the initial diagnostic exome analysis did not include validated genome-wide CNV calling, the *FBXO31* locus was subsequently reanalyzed using IGV.

### Segregation analysis

Segregation of the *FBXO31* deletion was assessed by PCR amplification and Sanger sequencing of a polymorphic amplicon in intron 5, located within the deleted interval. The amplicon contained two linked intronic polymorphisms that distinguished the amplifiable maternal and paternal alleles. Within this family, detection of both parental marker haplotypes was consistent with inheritance of two nondeleted chromosomes, whereas detection of only one parental haplotype indicated transmission of the deletion from the other parent. Failure to amplify the intron 5 amplicon in the proband was consistent with biallelic deletion of the assayed interval. Primer sequences, marker coordinates and electropherograms are provided in Fig. S1, Tables S2, S3.

### Deletion interval mapping and transcript consequence

The GRCh38 interval limits (intron 1 side, chr16:87,381,753; exon 9 side, chr16:87,327,440) were mapped against MANE Select exon and CDS coordinates for *FBXO31* (transcript NM_024735.5 / ENST00000311635; Ensembl release 116) retrieved from Ensembl. The F-box/SKP1-binding domain boundary (residues 64-110) was taken from UniProt (Q5XUX0). The Zn-beta motif (147-290) and a substrate-binding beta motif (291-539) were adopted from Li et al. (4).

### Population and in-silico variant analyses

*FBXO31* high-confidence predicted loss-of-function (pLoF) variants were obtained from gnomAD v4.1.1 on 24 July 2026, restricted to the MANE Select transcript (ENST00000311635), PASS calls and LOFTEE high-confidence with no flags (18, 19). Structural variants and copy-number variants overlapping *FBXO31* were retrieved from the gnomAD SV callset and from ClinVar; ClinVar copy-number variants were size-resolved on genome builds GRCh37/hg19 or GRCh38/hg38 and their genomic extent was compared with the patient’s deletion (20). Missense variant-effect predictions for all 10,241 possible *FBXO31* substitutions were obtained from AlphaMissense, and the two disease-associated missense variants, p.(Asp334Asn) and p.(Arg511Gln), were located within the prediction landscape (21).

### Use of large language models

The authors used ChatGPT (OpenAI, San Francisco, CA, USA) and Claude (Anthropic, San Francisco, CA, USA) to refine manuscript text and to revise analysis code. All model output was reviewed by the authors. Large language models were not used to interpret clinical data. The authors take full responsibility for the content of this article.

## Results

### Clinical report

We describe a female patient who was born with a normal birth weight (3200 g, 26th percentile) after an uneventful pregnancy to consanguineous parents (first cousins; Fig. 2a). Global developmental delay (GDD) (HP:0001263) was evident from early childhood (22). Both speech and motor milestones were delayed (delayed speech and language development, HP:0000750; delayed gross motor development, HP:0002194). Speech remained limited to single words, and at 5-9 years of age the Reynell Developmental Language Scales indicated comprehension at an approximately 1-year developmental level. She has persistent severe expressive and receptive language delay and currently uses 20-30 words and gestures.

From preschool age she exhibited behavioral problems (HP:0000708), including episodes of frustration-induced screaming and occasional aggression toward others, which improved over time but still occur intermittently.

At 5-9 years of age she was diagnosed with short stature (HP:0004322), microcephaly (HP:0000252) and low body weight (HP:0004325). By 10-14 years of age she showed partial catch-up growth (height 7th, weight 31st, head circumference 5th percentile) and no longer met criteria for microcephaly. Dysmorphic features included up-slanting palpebral fissures (HP:0000582), smooth philtrum (HP:0000319), thin upper-lip vermilion (HP:0000219), broad nasal tip (HP:0000455) and abnormal ear morphology (HP:0031703). Metabolic investigations were unremarkable apart from a transient alanine and alanine/lysine ratio elevation; EEG showed abnormal background activity. Cognitive testing indicated moderate intellectual disability (IQ <50; HP:0002342). Her clinical features are summarized alongside previously reported individuals with pathogenic *FBXO31* variants in Table 1.

**Table 1.** Clinical and genetic comparison of individuals with recessive *FBXO31*-associated neurodevelopmental disorder and dominant *FBXO31*-related Kruer syndrome. Clinical features of previously reported individuals with biallelic FBXO31 loss of function (Mir et al.; Moudi et al.) are compared with the present patient carrying a homozygous intragenic *FBXO31* deletion. Individuals with the recurrent heterozygous p.(Asp334Asn) gain-of-function variant reported by Jin et al., Dzinovic et al. and Galaz-Montoya et al. are included to illustrate the phenotypic distinction between recessive *FBXO31* loss of function and dominant Kruer syndrome.

| Category | Mir et al., 2014 (Pakistan) | Moudi et al., 2022 (Iran) | Present study | Galaz-Montoya et al., 2026 (including Jin et al., 2020 and Dzinovic et al., 2021) |
| --- | --- | --- | --- | --- |
| *Individuals included* | *6 affected individuals* | *2 affected siblings* | *1 affected individual (II-6)* | *7 unrelated individuals with recurrent de novo p.(Asp334Asn)* |
| **Inheritance / mechanism** | Autosomal recessive; predicted loss of function with reduced transcript and reduced protein abundance | Autosomal recessive; molecular effect unresolved | Autosomal recessive; predicted loss of function | Autosomal dominant; neomorphic gain of function |
| **Motor development** |  |  |  |  |
| **Motor skills** | Not reported | Not reported | Delayed walking | Motor developmental impairment in all individuals |
| **Speech development** |  |  |  |  |
| **Communication** | Not reported | Speech impairment in both siblings | First words delayed; 20-30 single words and gestures; no phrase speech | Language disorder or speech impairment in 7/7 |
| **Cognitive development** |  |  |  |  |
| **Development / Intelligence** | Mild to moderate ID (5/5 assessed) | Moderate ID in both siblings; GDD not clearly reported | GDD; moderate ID | GDD/ID in 7/7 |
| **Behavior** | Not reported | Not reported | Irritability; aggressive behavior | Stereotypies 6/7; anxiety 3/7; ADHD and aggressive behavior 2/7 each; ASD 1/7; self-injury 1/5 assessed; happy demeanor 5/5 assessed |
| **Neurology** |  |  |  |  |
| **Neurological findings** | No abnormalities on neurological examination (5/5 assessed) | Not reported | Abnormal EEG background activity; no other neurological diagnosis made | Cerebral palsy 5/7; diplegic spasticity 6/7; hypotonia 5/7; dystonia 2/6 assessed; hereditary spastic paraplegia 1/7; abnormal EEG in 1/3 assessed |
| **Brain imaging** | MRI reported in 2/5 assessed individuals; both normal | Normal MRI in both siblings | Not performed | Abnormal MRI in 6/7: thin/hypoplastic corpus callosum 4/7, ventriculomegaly 3/7, white-matter abnormalities 3/7, and gray-matter heterotopia 1/7 |
| **Seizures** | No history of seizures (5/5 assessed) | Seizures at 3 months | No seizures reported | Not reported |
| **Phenotype** |  |  |  |  |
| **Dysmorphic features** | Coarse facial features including broad nasal bridge, fleshy nares, and thick prominent lips | Long face, prominent supraorbital ridges, prominent jaw, fleshy nares, thick eyebrows, and thick prominent lips; no coarse facial features | Upslanted palpebral fissures, smooth philtrum, thin upper-lip vermillion, broad nasal tip, and abnormal ear morphology | Subtle shared facial gestalt: long face, broad/prominent forehead, sparse eyebrows, thin upper lip, and retrognathia in 7/7; strabismus in 5/7 |
| **Anthropometric data** | Not reported | Mild microcephaly in both siblings; weight and height not reported | Weight 31st percentile; height 7th percentile; head circumference 5th percentile at 10-14 years (microcephaly at 5-9 years, with subsequent partial catch-up) | Postnatal data not reported |
| **Genetics** |  |  |  |  |
| **FBXO31 variant** | c.847_852delinsA p.(Cys283Asnfs*81) | c.1532G>A p.(Arg511Gln) | Homozygous intragenic deletion of exons 2-8 and the coding portion of exon 9 | c.1000G>A p.(Asp334Asn) |
| **Zygosity** | Homozygous | Homozygous | Homozygous | Heterozygous |
| **Consanguinity** | Yes | Yes | Yes (parents are first cousins) | Not reported |

### A homozygous deletion removes the FBXO31 substrate-recognition region

Conventional karyotyping and *FMR1* testing were unremarkable, and initial exome sequence-variant analysis identified no alternative pathogenic variant. Chromosomal microarray subsequently identified a homozygous deletion encompassing the coding regions of exons 2-9 in *FBXO31* (Fig. S4). The array also detected three long contiguous stretches of homozygosity larger than 10 Mb, on 2p24.1-p22.3, 11q22.1-q23.1 and Xp11.4-p11.1 consistent with parental relatedness. Since copy-number detection was not routinely implemented in the diagnostic exome pipeline at the time, exome sequencing data were reanalyzed and showed concordant loss of coverage across the deleted *FBXO31* exons, mapping the deleted interval to chr16:87,327,440-87,381,753 (Fig. 1a; Fig. S5). Under Ensembl release 116, the interval also partially overlaps an uncharacterized protein-coding annotation (*ENSG00000131152*) and overlaps two lncRNA annotations (*ENSG00000269901* and *ENSG00000270082*). *FBXO31* was prioritized as the leading candidate because the deletion removes most of its coding sequence and because prior human genetic and mechanistic evidence provides an independent *FBXO31* disease rationale.

**Figure 1.**
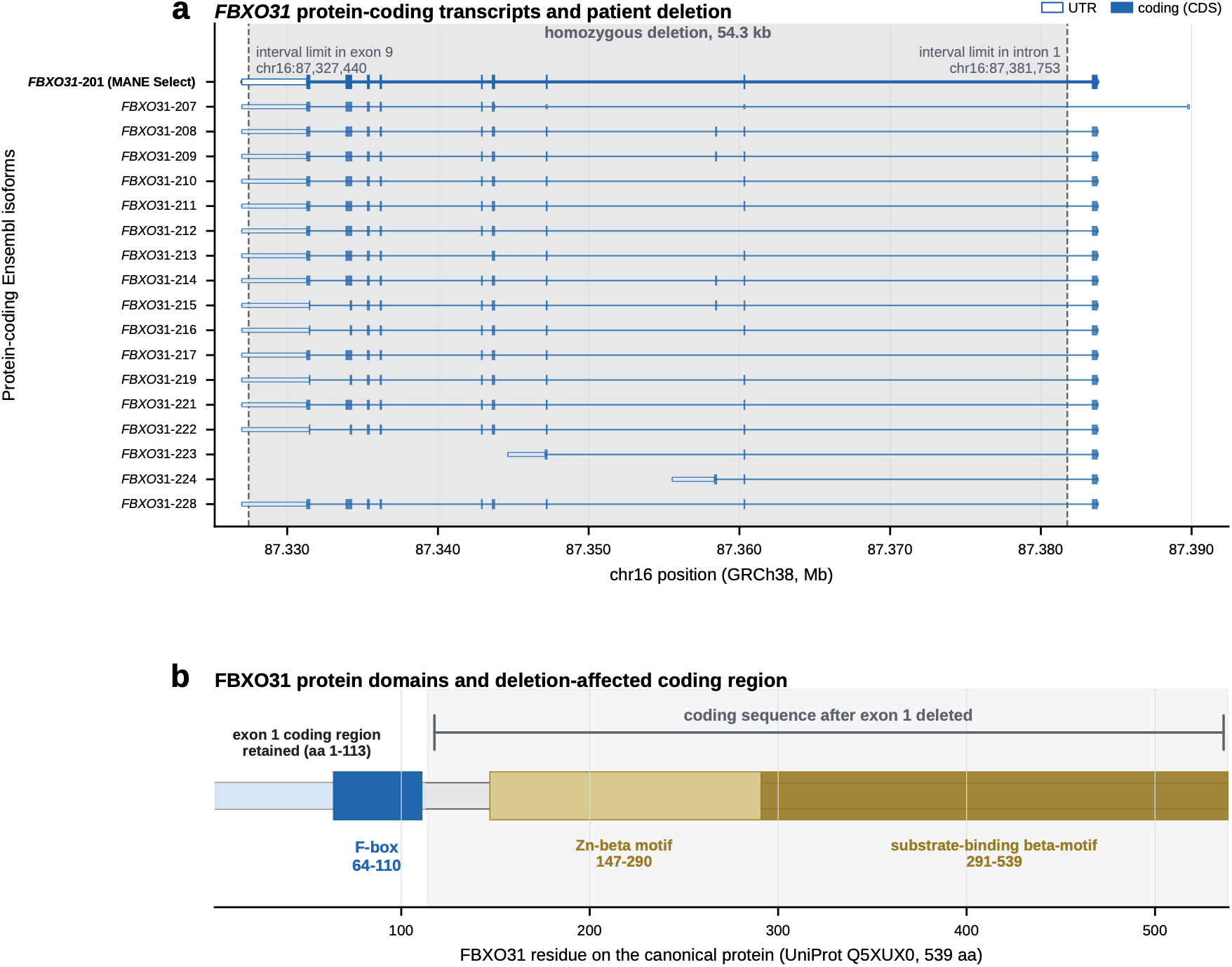
The homozygous *FBXO31* deletion removes the substrate-recognition region across annotated protein-coding transcripts. (a) The patient’s ~54 kb homozygous deletion (GRCh38 chr16:87,327,440-87,381,753) shown against *FBXO31* transcripts; the gene is on the minus strand. The deletion removes exons 2-8 and the coding part of exon 9 while retaining exon 1, disrupting the coding sequence of each of the 18 annotated protein-coding transcripts. (b) Consequence at the protein level: the retained exon 1 encodes the SKP1-binding F-box (residues 64-110), whereas the deleted exons encode the entire C-terminal substrate-recognition region including a Zn-beta motif (147-290) and a substrate-binding beta motif (291-539).

The retained exon 1 contains the coding sequence for residues 1-113, including the SKP1-binding F-box domain, whereas the deletion removes the coding sequence for the downstream C-terminal substrate-recognition region (Fig. 1b). In the MANE Select transcript, the interval removes 1,280 of 1,620 coding base pairs (79.0%). It also overlaps the coding sequence of all 18 protein-coding *FBXO31* transcripts retrieved from Ensembl release 116 (Fig. 1a). The deletion is therefore predicted to result in severe loss of function, through loss of the substrate-recognition module and/or degradation of any truncated transcript.

### Segregation and population data are compatible with a recessive loss-of-function mechanism

Segregation analysis confirmed that both parents are heterozygous carriers, each transmitting one deleted allele. Inheritance patterns were assessed using an amplicon located within the deleted interval that contained closely spaced intronic polymorphisms distinguishing the maternal and paternal nondeleted chromosomes (Tables S2, S3). The patient showed no amplification, consistent with biallelic deletion. In the siblings, transmission of a deletion allele was inferred from the absence of parental marker haplotypes (Fig. S1). Among the five unaffected siblings, two were homozygous for the wild-type allele, one carried the maternal deletion and two carried the paternal deletion; no neurodevelopmental abnormalities were present in any heterozygous carrier (Fig. 2a). This pattern (affected only in the homozygous state, unaffected in heterozygous carriers) is consistent with autosomal-recessive inheritance.

**Figure 2.**
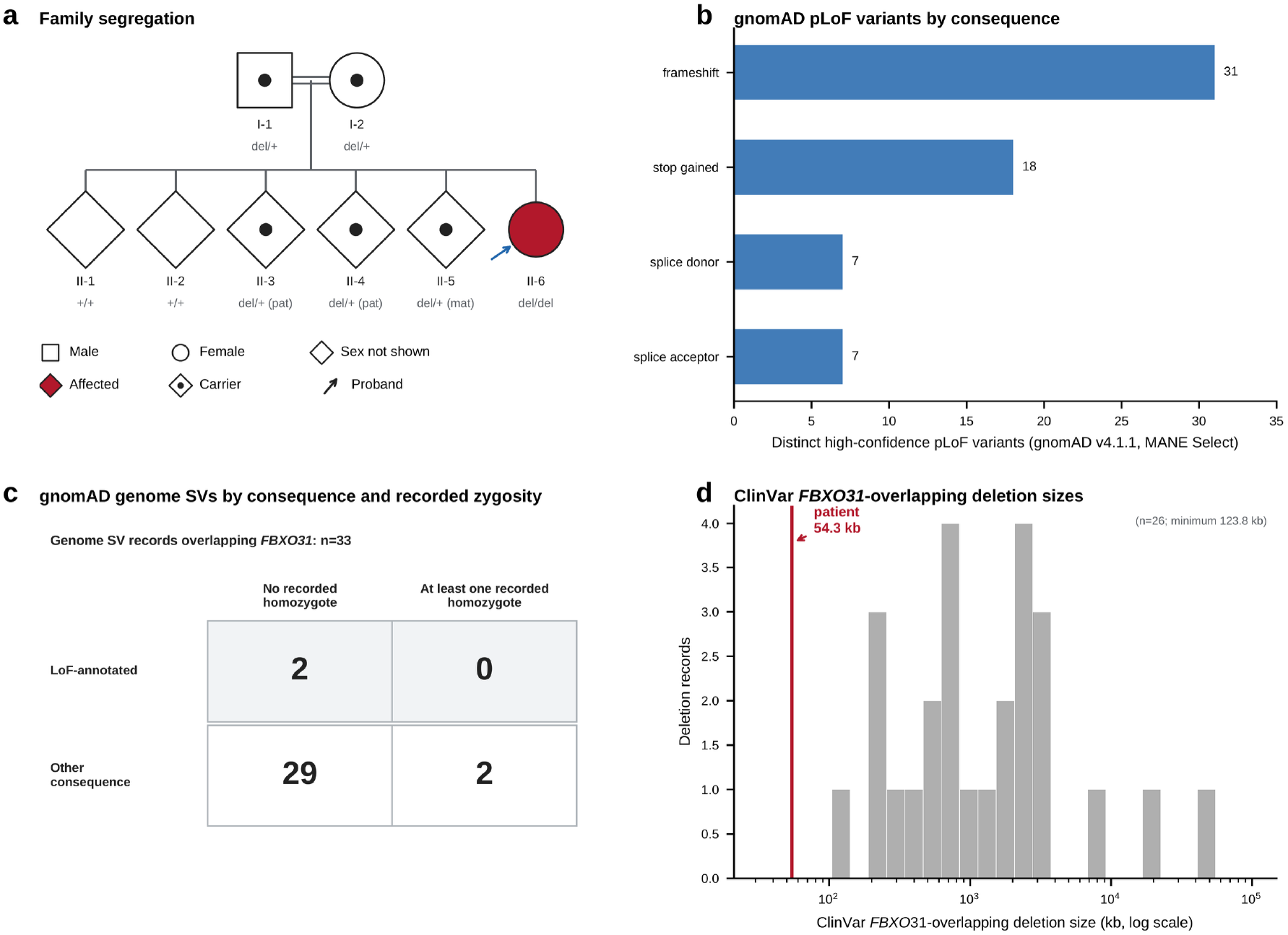
Segregation and population evidence for a recessive loss-of-function mechanism. (a) Pedigree of the consanguineous family: the affected proband is homozygous for the *FBXO31* deletion; both parents and three of five genotyped, unaffected siblings are heterozygous carriers, and two siblings are homozygous for the wild-type allele. (b) PASS, LOFTEE high-confidence pLoF variants without flags affecting the MANE Select *FBXO31* transcript in gnomAD v4.1.1: 63 distinct variants with a summed alternate allele count of 89 and no recorded same-variant homozygotes. (c) Consequence annotation and recorded homozygotes among 33 PASS gnomAD genome SVs overlapping *FBXO31*. The two LoF-annotated SVs had no recorded homozygotes. Two non-LoF-annotated SVs each had a recorded homozygote: a 1.6-kb intronic deletion with no MANE coding-sequence overlap and a 58.3-kb partial-gene duplication overlapping coding sequence. Neither represented a homozygous coding-loss SV comparable to the patient’s allele. Eleven additional exome CNV records were not included in this comparison because an equivalent homozygote-count field was unavailable. (d) Sizes of 26 *FBXO31*-overlapping ClinVar deletion/loss records with resolvable spans after exclusion of two drug-response records. All were multigene events and were larger than the patient’s focal ~54 kb deletion (minimum 123.8 kb; median 1.28 Mb).

Population variation is consistent with this interpretation. In gnomAD v4.1.1, 63 distinct high-confidence pLoF single nucleotide variants (SNVs) or small indels are reported for the MANE Select *FBXO31* transcript, none of which were observed in the homozygous state (Fig. 2b, Table S7). Likewise, no LoF-annotated structural variant (SV) overlapping *FBXO31* in gnomAD had a recorded homozygote (Fig. 2c, Table S7). The only two SVs with recorded homozygotes were an intronic deletion and a partial-gene duplication overlapping coding sequence, neither of which was annotated as a loss-of-function event. Thus, heterozygous predicted loss-of-function variation is present in the population, whereas no homozygous coding-loss equivalent of the patient deletion was identified.

To complement the population analysis, we examined whether clinically reported deletions provided evidence for a phenotype specifically attributable to heterozygous *FBXO31* loss. No comparable gene-focused deletion was identified in ClinVar (23). All 26 *FBXO31*-overlapping deletion/loss records with resolvable spans were larger than the patient interval (range 123.8 kb-43.5 Mb) and were multigene events (Fig. 2d, Table S7). Although these events also disrupt *FBXO31*, their multigene context precludes attributing their clinical interpretations specifically to *FBXO31*.

## Discussion

We describe an individual with moderate intellectual disability, developmental delay and dysmorphic features who carries a homozygous, focal intragenic deletion of *FBXO31*. To our knowledge, this is the first reported homozygous *FBXO31* deletion. The allele is particularly informative because its predicted consequence is independent of transcript fate: it removes exons 2-8 and the coding part of exon 9, leaving only exon 1 intact, so that even a residual transcript escaping nonsense-mediated decay could encode at most a short N-terminal fragment lacking the substrate-recognition region required for FBXO31-mediated ubiquitination (Fig. 1b). Together with the two previously reported recessive families, this allele provides human genetic evidence that biallelic *FBXO31* loss can underlie a recessive neurodevelopmental disorder.

Previous reports have linked homozygous *FBXO31* variants to neurodevelopmental disease, but the underlying mechanisms remained incompletely resolved. Mir et al. described a homozygous deletion-insertion, c.847_852delinsA p.(Cys283Asnfs*81), in a consanguineous family with intellectual disability and shared coarse facial features (16). Patient-derived lymphoblasts showed an approximately threefold reduction in *FBXO31* mRNA, consistent with partial nonsense-mediated decay, but reduction of protein levels was modest (approximately 1.4-fold), limiting conclusions about the extent of functional loss. Moudi et al. reported a homozygous p.(Arg511Gln) missense variant in two affected siblings, while heterozygous relatives were unaffected, but without functional validation (15). The AlphaMissense score for p.(Arg511Gln) supports a functional impact for this substitution but does not establish loss of function (Fig. S6).

Neither population nor clinical variation databases argue against a recessive mechanism, and neither provides positive evidence for haploinsufficiency. Heterozygous pLoF variants are present in gnomAD, whereas no homozygous pLoF variant overlapping *FBXO31* was identified. Because the individual variants are rare, however, the absence of homozygotes is expected and cannot be interpreted as evidence of intolerance to biallelic loss. ClinVar likewise provides no gene-specific evidence that heterozygous deletion of *FBXO31* alone causes disease: all 26 size-resolved *FBXO31*-overlapping copy-number deletions are larger multigene events, precluding attribution of their clinical manifestations specifically to *FBXO31* (Fig. 2d). The present deletion is distinct in that both reported interval limits lie within *FBXO31*.

Recent work provides a mechanistic rationale for *FBXO31* deletion. Muhar et al. showed that FBXO31 knockout abolished CTAP clearance, while knockdown induced a transcriptional response in neural progenitors that became more pronounced in mature neurons (6). The same study showed that c.1000G>A p.(Asp334Asn) is not a simple loss-of-function allele, but redirects FBXO31 towards non-CTAP substrates.

Together with our observations, this supports a mechanism- and zygosity-specific model of *FBXO31*-associated disease (Fig. 3). The recurrent heterozygous p.(Asp334Asn) variant acts through a dominant gain-of-function mechanism and causes Kruer syndrome (8). Heterozygous loss of function alone is likely not sufficient to cause disease, whereas biallelic loss of function would abolish canonical FBXO31 activity without conferring neomorphic substrate targeting. Clinically, these mechanisms correspond to two distinct disorders (Table 1). The dominant p.(Asp334Asn) phenotype is defined by motor involvement, with cerebral palsy accompanying intellectual disability. Biallelic loss of function is instead intellectual-disability-predominant, without symptoms of cerebral palsy.

**Figure 3.**
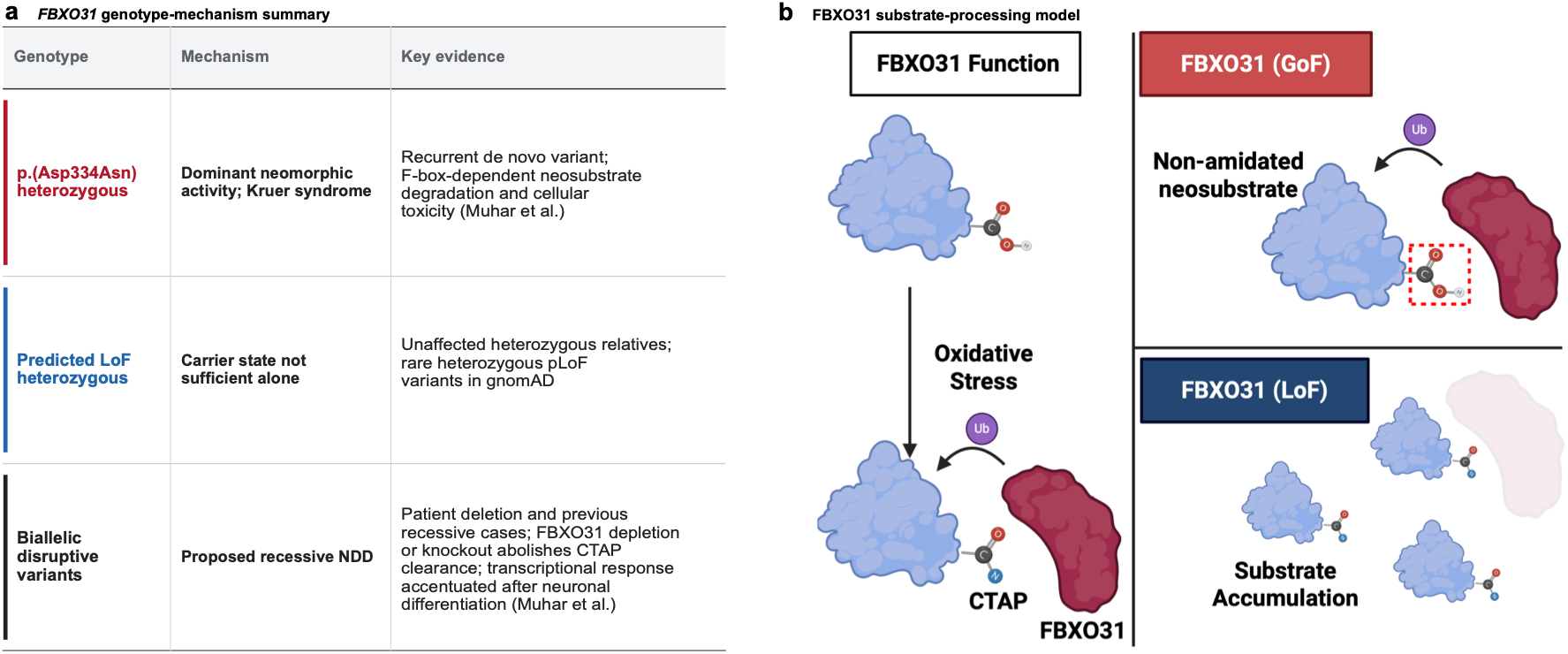
A proposed *FBXO31* allelic series spanning gain and loss of function. (a) The three genotype classes and their consequences: heterozygous p.(Asp334Asn) (dominant gain-of-function/neomorphic activity leading to Kruer syndrome); heterozygous loss of function (insufficient alone, based on unaffected carriers and gnomAD heterozygous pLoF); and biallelic loss of function (recessive neurodevelopmental disorder, supported by the homozygous patient deletion). (b) Proposed functional model: normal *FBXO31* clears CTAP-marked substrates via the SCF complex; the gain-of-function variant redirects the ligase toward aberrant substrates, whereas biallelic loss is predicted to impair CTAP-substrate clearance.

Two considerations qualify the interpretation of our case. First, because exon 1 is retained, a residual N-terminal fragment could in principle occupy the SCF core without delivering substrates. A dominant-negative effect is nevertheless unlikely, as substrate receptors that cannot bind substrates are preferentially released from the CUL1-RBX1 core (24-27), and all five heterozygous carriers in this family are unaffected. Second, the deleted interval is not annotation-exclusive: it also intersects one uncharacterized protein-coding locus and two lncRNAs, and a contribution from these co-deleted annotations cannot be formally excluded. *FBXO31* was nevertheless prioritized because its coding sequence is directly disrupted and because independent human genetic and mechanistic evidence implicates the gene in neurodevelopmental disease.

Our findings strengthen the *FBXO31*-disease relationship, currently listed as provisional in OMIM (#615979), and support biallelic *FBXO31* loss of function as a cause of autosomal-recessive neurodevelopmental disease, mechanistically distinct from the dominant gain-of-function disorder caused by p.(Asp334Asn).

## Data Availability

All data produced in the present study are available upon reasonable request to the authors

## Data availability

The patient-level genetic data generated in this study contain potentially identifiable information and cannot be made publicly available for reasons of patient privacy. Access to relevant data may be requested from the corresponding author, subject to applicable ethical and institutional approvals.

## Code availability

The analysis code used to generate the population, structural-variant, ClinVar and AlphaMissense summaries and the figure-level counts reported in this study is available from the corresponding author upon reasonable request.

## Acknowledgements

BioRender was used to create Fig. 3b (publication license obtained).

## Author contributions

JB: data analysis, study conception, manuscript drafting and figure preparation. BW: genetic diagnostics and Sanger sequencing. SH: manuscript drafting, figure preparation. JM: manuscript drafting, figure preparation. KO: genetic diagnostics. KH: genetic diagnostics. SSc (Sebastian Schröder): clinical evaluation of our patient. BS: clinical evaluation of our patient. MH: clinical evaluation of our patient, manuscript revision and supervision. SSa (Sebastian Sailer): figure preparation, supervision, manuscript revision, clinical and genetic diagnostics.

## Ethical approval

Ethical approval was granted by the local ethics committee in Heidelberg (S-632/2023). Written informed consent for publication was obtained from all participants or their legal guardians.

