## Supplemental Information for "Recessive loss-of-function and dominant gain-of-function variants in *FBXO31* are associated with distinct neurodevelopmental disorders"

**Supplementary Information**

**Supplementary Figure S1**

**(a) PCR across a polymorphic amplicon in intron 5, within the deleted interval**

| **Lane** | **Gel label** | **Individual** | **Genotype** | **Product** |
| --- | --- | --- | --- | --- |
| 1 | B | II-6 (proband) | del/del | Absent |
| 2 | C | II-1 | +/+ | Present |
| 3 | D | II-2 | +/+ | Present |
| 4 | E | II-3 | del/+ | Present |
| 5 | F | I-2 (mother) | del/+ | Present |
| 6 | G | I-1 (father) | del/+ | Present |
| 7 | neg | No-template control | — | Absent |

**(b) Sanger electropherograms of the intron 5 amplicon**

| 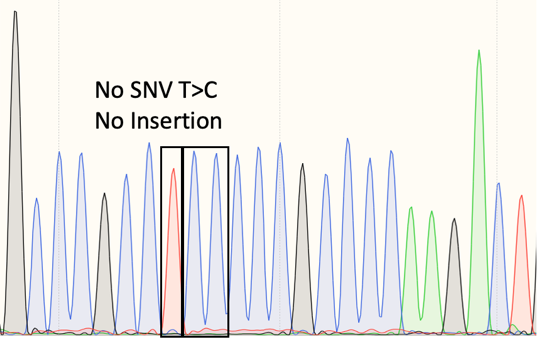  **I-1 Father,** del/+  Reference haplotype only: no c.T>C, no insertion | 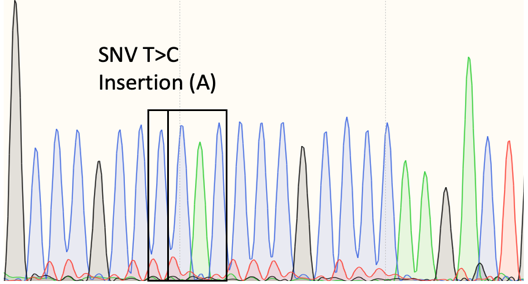  **I-2** Mother, del/+  Marker haplotype only: c.T>C and insertion (A) |
| --- | --- |
| 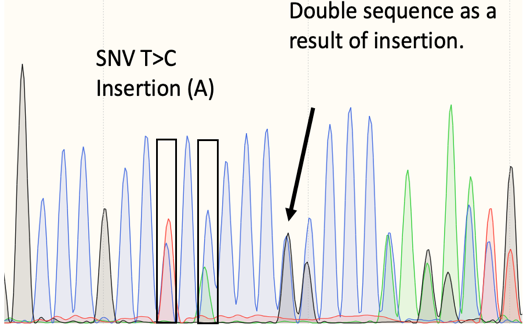  **II-1**, +/+  Both parental haplotypes; double sequence after the insertion | 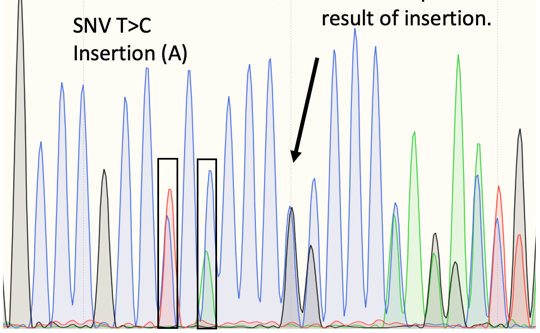  **II-2**, +/+  Both parental haplotypes; double sequence after the insertion |
| 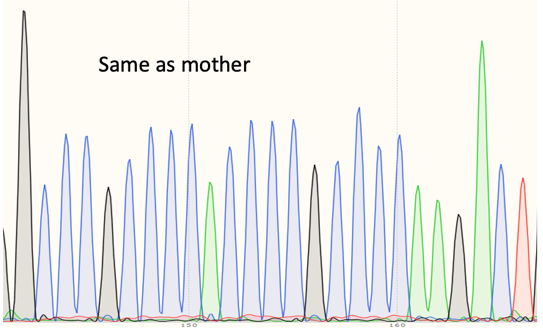  **II-3**, del/+ (pat)  Maternal haplotype only | 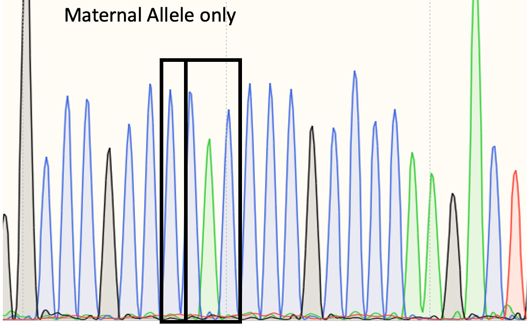  **II-4**, del/+ (pat)  Maternal haplotype only |
| 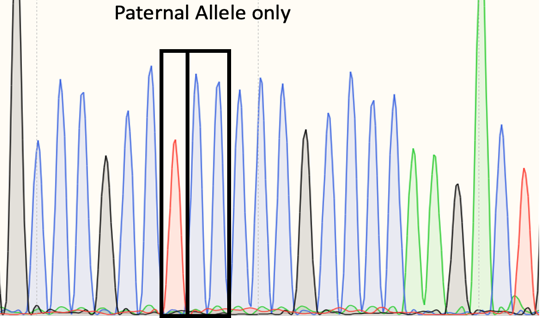  **II-5**, del/+ (mat)  Paternal haplotype only | **II-6** Proband, del/del  No PCR product obtained; no electropherogram. |

**Supplementary Figure S1.** Segregation of the *FBXO31* deletion. (a) Agarose gel of the intron 5 amplicon for the proband, siblings and parents, together with a no-template control. No product was obtained from the proband (lane 1), consistent with homozygous deletion of the assayed interval. (b) Electropherograms. The maternal non-deleted chromosome carries both linked markers (c.T>C and an inserted A); the paternal non-deleted chromosome carries neither. A child showing both haplotypes therefore inherited two non-deleted chromosomes, whereas a child showing only one parental haplotype inherited the deletion from the other parent. The sex of the unaffected siblings is not shown.

**Supplementary Table S2**

**Intron 5 marker variants used for segregation analysis.**

| **Property** | **Marker 1** | **Marker 2** |
| --- | --- | --- |
| dbSNP identifier | rs34400756 | rs11457751 |
| ClinGen Allele Registry | CA16532114 | CA286130733 |
| GRCh38 (NC_000016.10) | g.87338472A>G | g.87338470_87338471insT |
| GRCh37 (NC_000016.9) | g.87372078A>G | g.87372076_87372077insT |
| Location | *FBXO31* intron 5, within the deleted interval | *FBXO31* intron 5, 2 bp from marker 1 |

**Supplementary Table S3**

**PCR primers used for segregation analysis.**

| **Item** | **Value** |
| --- | --- |
| Target region | *FBXO31* intron 5 (GRCh38 chr16), within the homozygously deleted interval |
| Primer 1 | CCATTCCTTGTACCCAGGCT |
| Primer 2 | GCCGTCCCAAGCATGTTTAT |

**Supplementary Figure S4**

Chromosomal microarray of the index patient. Genomic DNA from peripheral blood leukocytes was hybridized onto the Affymetrix CytoScan HD array and analyzed with Chromosome Analysis Suite v4.2.1 (GRCh37 annotations). A homozygous deletion on chromosome 16 encompassing coding regions of exons 2-9 in *FBXO31* was detected. Three long contiguous stretches of homozygosity exceeding 10 Mb were identified on chromosomes 2p24.1-p22.3, 11q22.1-q23.1 and Xp11.4-p11.1, consistent with parental consanguinity.

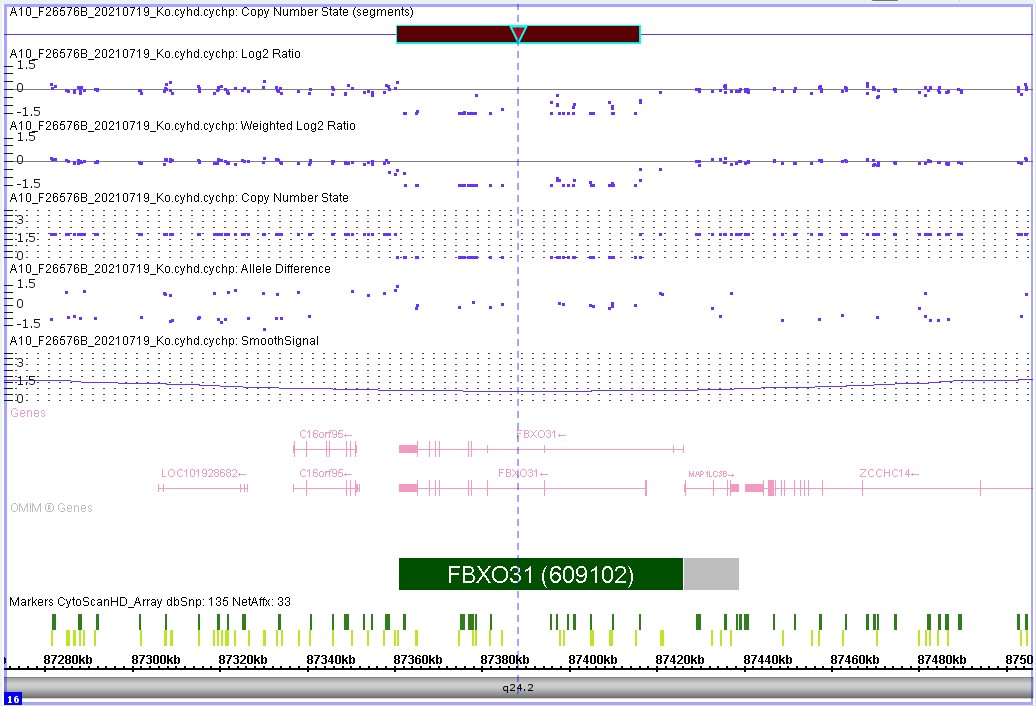

**Supplementary Figure S5**

Exome read-depth profile at the *FBXO31* locus. Exome sequencing data (SureSelect Human All Exon V7; NovaSeq 6000; GRCh38 alignment via the varfeed pipeline) were reanalyzed in IGV for copy-number inference. Loss of read depth is apparent across FBXO31 exons 2-8 and the coding portion of exon 9, concordant with the homozygous deletion identified by microarray and defining the interval chr16:87,327,440–87,381,753.

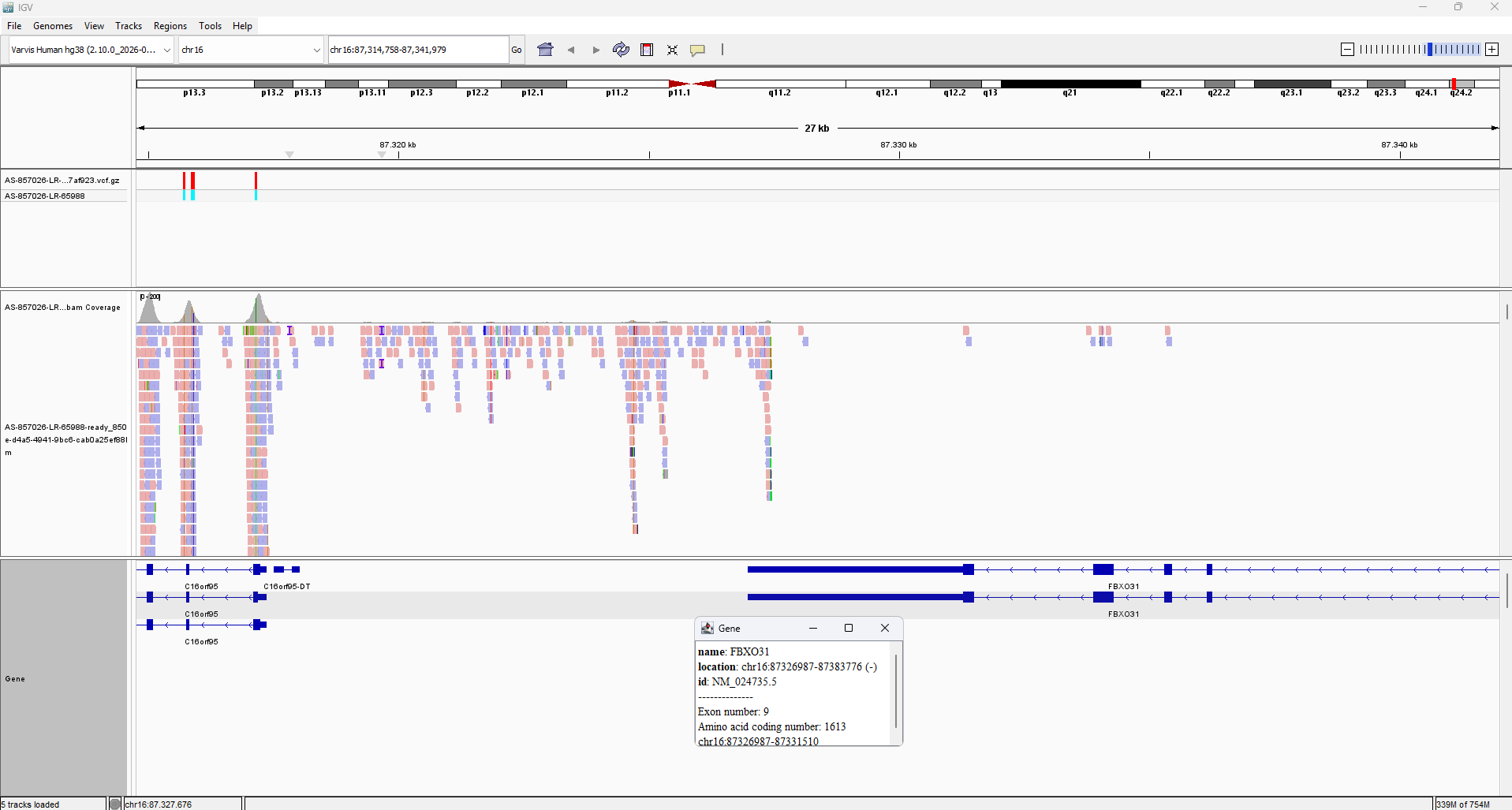

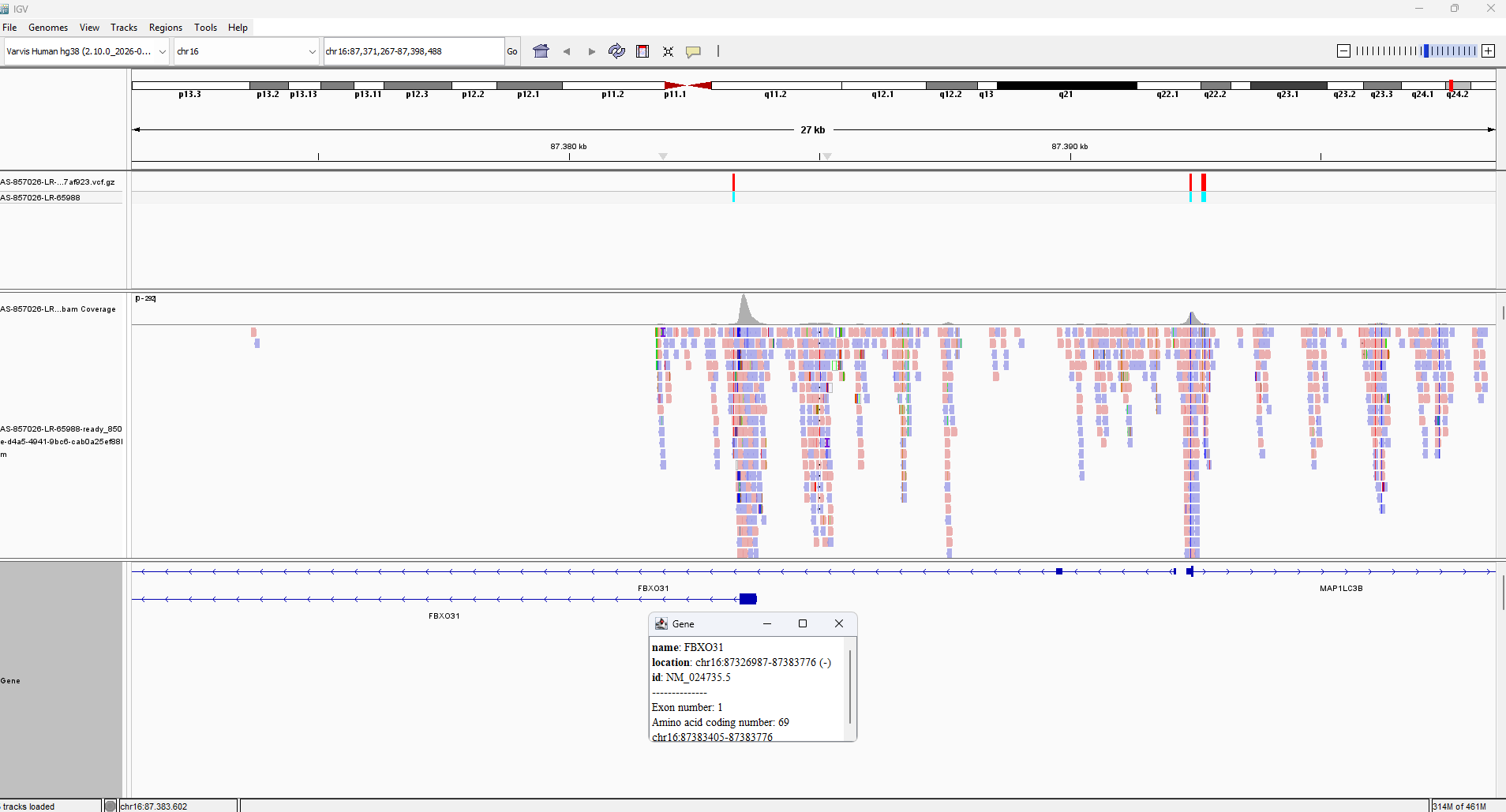

**Supplementary Figure S6**

AlphaMissense pathogenicity landscape across *FBXO31*. Per-residue mean AlphaMissense pathogenicity scores are plotted for all single amino acid substitutions across the 539-residue FBXO31 sequence. The dominant gain-of-function variant p.(Asp334Asn) (AM = 0.995; likely pathogenic) and the recessive missense p.(Arg511Gln) reported by Moudi et al. (AM = 0.701; likely pathogenic) are annotated.

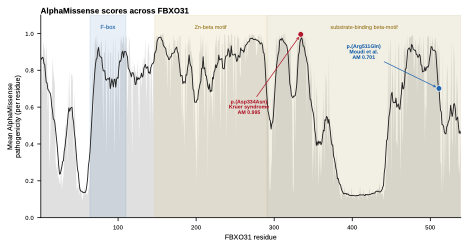
